# COVID-19 Hospitalization is More Frequent and Severe in Down Syndrome and Affects Patients a Decade Younger.

**DOI:** 10.1101/2020.05.26.20112748

**Authors:** Louise Malle, Cynthia Gao, Nicole Bouvier, Bethany Percha, Dusan Bogunovic

## Abstract

**Background:** Individuals with rare disorders, like Down syndrome (DS) are historically understudied. Currently, it is not known how COVID-19 pandemic affects individuals with DS. Herein, we report an analysis of individuals with DS who were hospitalized with COVID-19 in the Mount Sinai Health System in New York City, USA.

**Methods:** In this retrospective, single-center study of 4,615 patients hospitalized with COVID-19, we analyzed all patients with DS admitted in the Mount Sinai Health System. Hospitalization rates, clinical and outcomes were assessed.

**Findings:** Contrary to an expected number of one, we identified six patients with DS. We found that patients with DS are at an 8.9-fold higher risk of hospitalization with COVID-19 when compared to non-DS patients. Hospitalized DS individuals are on average 10 years younger than non-DS patients with COVID-19. Moreover, type 2 diabetes mellitus appears to be an important driver of this susceptibility to COVID-19. Finally, patients with DS have more severe outcomes than controls, and are more likely to progress to sepsis in particular.

**Interpretation:** We demonstrate that individuals with DS represent a higher risk population for COVID-19 compared to the general population and conclude that particular care should be taken for both the prevention and treatment of COVID-19 in these patients.

**Funding:** National Institute of Allergy and Infectious Diseases.

**Research in context:** *Evidence before this study:* We searched PubMed and Google Scholar on May 26, 2020, for articles describing the features of patients in Down syndrome infected with severe acute respiratory syndrome coronavirus 2 (SARS-CoV-2), using the search terms “SARS-CoV-2” or “COVID-19” and “Down syndrome” or “Trisomy 21.” We found only one case report describing a cluster of 4 cases of COVID-19 in a healthcare facility for patients with mental retardation.

*Added value of this study:* We compared the hospitalization rates of DS patients to over 4,500 individuals without DS, and we assessed comorbidities and outcomes of individuals with DS compared to age, race, and sex-matched controls hospitalized with COVID-19. To the best of our knowledge, we provide the first evidence that patients with DS with are at higher risk of hospitalization with COVID-19 and more severe disease progression than non-DS patients.

*Implications of all the available evidence:* We demonstrate that individuals with DS are a high-risk population for COVID-19 and suggest appropriate measures should be taken for both the prevention and treatment of COVID-19 in these patients.

## Introduction

Infection with the novel coronavirus severe acute respiratory syndrome-related coronavirus 2 (SARS-CoV-2) causes symptoms ranging from fever, cough and fatigue to severe lung injury and death. This disease, termed Coronavirus Disease-2019 (COVID-19), has caused a global pandemic of historic proportions. Although there are approximately a quarter million individuals with Down syndrome (DS) in the US alone, it is unknown how SARS-CoV-2 affects people with DS. Moreover, these individuals are known to be at higher risk of respiratory tract infections, particularly respiratory syncytial virus and influenza infections.^1,2^ Therefore, timely, evidence-based analyses of SARS-CoV-2 infection in DS are warranted.

At the time of our study, 4,615 total patients had been hospitalized in the Mount Sinai Health System due to COVID-19. In contrast to the expected single patent, we identified six individuals with DS. In this study, we aim to better understand the risk of COVID-19 hospitalization in individuals with DS, as well as the disease progression in these patients.

## Methods

### Study design and participants

For this retrospective, single-center study, we recruited patients admitted in the Mount Sinai Health System (New York City, USA) from March 1 to April 24, 2020 with a diagnosis of COVID-19 confirmed by RT-PCR. All patients with DS who were diagnosed with COVID-19 were included (n=6). Five randomly sampled age, race, and sex-matched controls were included for each DS patient (30 controls in total). DS patients for whom ethnicity was not specified in the electronic medical record were matched with controls of unspecified ethnicity.

### Procedures

We obtained demographic data, vitals, medications, laboratory values, diagnoses, and outcome data from patients’ electronic medical records.

### Statistical analysis

Statistical analysis was done using R. Confidence intervals for hospitalization rates (Table 1) were obtained using a 1-sample proportions test with continuity correction with the prop.test function. Comparison of comorbidities and complications between DS patients and controls were obtained by Fisher’s exact test (Table 2). Competing risk analysis and comparison of cumulative incidence curves was used for health outcomes (Table 2, Supplementary Figure 1C). Unpaired t-tests were used for analysis of laboratory values (Supplementary Figures 1A, 1B).

**Table 1.** Estimated COVID-19 hospitalization rates in the Mount Sinai Health System in DS and non-DS patients.

|  | All patients |  |  | Down syndrome |  |  |
| --- | --- | --- | --- | --- | --- | --- |
| Age (years) | Hospitalized<br>for COVID-19 | Seen at Mt Sinai in<br>5 years* | Per 1,000 | Hospitalized<br>for COVID-19 | Seen at Mt Sinai<br>in 5 years* | Per 1,000 [95 CI] |
| 30-34 | 128 | 243,514 | 0.53 | 1 | 66 | 15.15 [0.79-92.6] |
| 50-54 | 316 | 189,000 | 1.67 | 2 | 72 | 27.78 [4.83-105.8] |
| 55-59 | 438 | 191,691 | 2.84 | 2 | 71 | 28.17 [4.83-132.3] |
| 60-64 | 533 | 179,814 | 2.96 | 1 | 45 | 22.22 [1.16-132.3] |
| 30-64 | 1,972 | 1,361,679 | 1.45 | 6 | 468 | 12.8 [5.22-29.1] |

\*unique patients who had an encounter in the Mount Sinai Health System between 11/01/2015 and 04/24/2020.

**Table 2.**
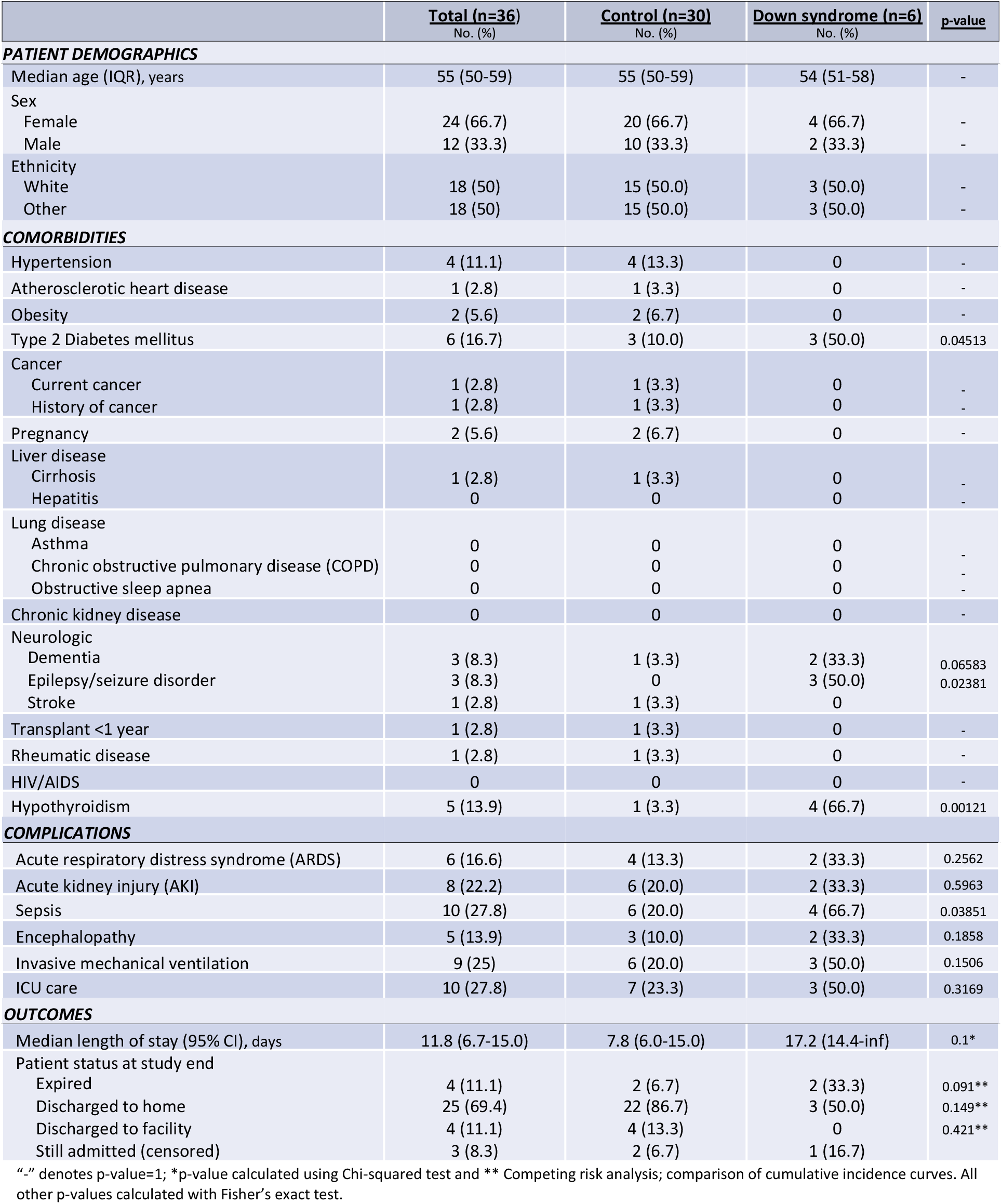
Demographics, comorbidities, disease complications, and outcomes of Down syndrome patients and age, sex, and ethnicity-matched controls hospitalized for COVID-19.

|  | **Total (n=36)**<br>No. (%) | **Control (n=30)**<br>No. (%) | **Down syndrome (n=6)**<br>No. (%) | **p-value** |
| --- | --- | --- | --- | --- |
| **PATIENT DEMOGRAPHICS** |  |  |  |  |
| Median age (IQR), years | 55 (50-59) | 55 (50-59) | 54 (51-58) | - |
| Sex |  |  |  |  |
| Female | 24 (66.7) | 20 (66.7) | 4 (66.7) | - |
| Male | 12 (33.3) | 10 (33.3) | 2 (33.3) | - |
| Ethnicity |  |  |  |  |
| White | 18 (50) | 15 (50.0) | 3 (50.0) | - |
| Other | 18 (50) | 15 (50.0) | 3 (50.0) | - |
| **COMORBIDITIES** |  |  |  |  |
| Hypertension | 4 (11.1) | 4 (13.3) | 0 | - |
| Atherosclerotic heart disease | 1 (2.8) | 1 (3.3) | 0 | - |
| Obesity | 2 (5.6) | 2 (6.7) | 0 | - |
| Type 2 Diabetes mellitus | 6 (16.7) | 3 (10.0) | 3 (50.0) | 0.04513 |
| Cancer |  |  |  |  |
| Current cancer | 1 (2.8) | 1 (3.3) | 0 | - |
| History of cancer | 1 (2.8) | 1 (3.3) | 0 | - |
| Pregnancy | 2 (5.6) | 2 (6.7) | 0 | - |
| Liver disease |  |  |  |  |
| Cirrhosis | 1 (2.8) | 1 (3.3) | 0 | - |
| Hepatitis | 0 | 0 | 0 | - |
| Lung disease |  |  |  |  |
| Asthma | 0 | 0 | 0 | - |
| Chronic obstructive pulmonary disease (COPD) | 0 | 0 | 0 | - |
| Obstructive sleep apnea | 0 | 0 | 0 | - |
| Chronic kidney disease | 0 | 0 | 0 | - |
| Neurologic |  |  |  |  |
| Dementia | 3 (8.3) | 1 (3.3) | 2 (33.3) | 0.06583 |
| Epilepsy/seizure disorder | 3 (8.3) | 0 | 3 (50.0) | 0.02381 |
| Stroke | 1 (2.8) | 1 (3.3) | 0 | - |
| Transplant <1 year | 1 (2.8) | 1 (3.3) | 0 | - |
| Rheumatic disease | 1 (2.8) | 1 (3.3) | 0 | - |
| HIV/AIDS | 0 | 0 | 0 | - |
| Hypothyroidism | 5 (13.9) | 1 (3.3) | 4 (66.7) | 0.00121 |
| **COMPLICATIONS** |  |  |  |  |
| Acute respiratory distress syndrome (ARDS) | 6 (16.6) | 4 (13.3) | 2 (33.3) | 0.2562 |
| Acute kidney injury (AKI) | 8 (22.2) | 6 (20.0) | 2 (33.3) | 0.5963 |
| Sepsis | 10 (27.8) | 6 (20.0) | 4 (66.7) | 0.03851 |
| Encephalopathy | 5 (13.9) | 3 (10.0) | 2 (33.3) | 0.1858 |
| Invasive mechanical ventilation | 9 (25) | 6 (20.0) | 3 (50.0) | 0.1506 |
| ICU care | 10 (27.8) | 7 (23.3) | 3 (50.0) | 0.3169 |
| **OUTCOMES** |  |  |  |  |
| Median length of stay (95% CI), days | 11.8 (6.7-15.0) | 7.8 (6.0-15.0) | 17.2 (14.4-inf) | 0.1* |
| Patient status at study end |  |  |  |  |
| Expired | 4 (11.1) | 2 (6.7) | 2 (33.3) | 0.091** |
| Discharged to home | 25 (69.4) | 22 (86.7) | 3 (50.0) | 0.149** |
| Discharged to facility | 4 (11.1) | 4 (13.3) | 0 | 0.421** |
| Still admitted (censored) | 3 (8.3) | 2 (6.7) | 1 (16.7) |  |

"-" denotes p-value=1; \*p-value calculated using Chi-squared test and \*\* Competing risk analysis; comparison of cumulative incidence curves. All other p-values calculated with Fisher's exact test.

## Results

In total, 4,615 total patients were hospitalized for COVID-19 in the Mount Sinai Health System in the 55 days beginning on March 1, 2020. Over the last five years, this facility has cared for over 2.6 million individuals, 1,148 of whom had DS. Assuming that the COVID-19 patients were a representative sample of the Mount Sinai patient population, we would therefore have expected approximately two individuals with DS to be among the patients hospitalized for COVID-19. Furthermore, as the mean age of individuals with DS is significantly lower than that of the general population,^3^ the expected number of patients hospitalized is closer to one. Contrary to expectations, six of the patients hospitalized with COVID-19 had DS. We found that the incidence of hospitalization in the DS population was 8.9 times higher (CI: 4.00-20.0) than that in the general population, for individuals 30-64 years old (Table 1).

The median age of patients with DS hospitalized with COVID-19 was 54 years (interquartile range [IQR]: 51-58 years) (Table 2). These patients were thus significantly younger than the rest of the population hospitalized at Mount Sinai (median: 66 years, IQR: 55-78) and in previous studies (median: 63 years, IQR: 52-75).^4^ These findings suggest that DS patients are at an increased risk of being hospitalized for COVID-19 at a significantly younger age.

We then compared the hospitalized individuals with DS to a control group composed of randomly sampled non-DS patients hospitalized with COVID-19, with five such patients matched for sex, age, and ethnicity for each patient with DS (30 controls in total). DS patients for whom ethnicity was not specified in the electronic medical record were matched with controls of unspecified ethnicity. The most common underlying conditions in the control group were hypertension (4, 13.3%), diabetes (3, 10%), and obesity (2, 6.7%). By contrast, hypothyroidism (4, 66.7%), type 2 diabetes (3, 50%), epilepsy (3, 50%), and dementia (2, 33.3%) were the only comorbidities found in the DS group (Table 2). This is consistent with the higher incidence of hypothyroidism, epilepsy, and dementia in the DS population.^5-7^ Moreover, it is estimated that patients with DS have a 3-4-fold increase in type I diabetes,^8^ although less is known about the incidence of type 2 diabetes in this population. Hypothyroidism is unlikely to contribute to COVID-19 given that it is well controlled pharmacologically. On the other hand, diabetes, which reached statistical significance despite the limited number of patients (p<0.05), is a likely underlying susceptibility factor for COVID-19 hospitalization and severity, as reported recently.^4,9^ Neurological disorders were also shown to increase the risk of COVID-19 mortality in one study, although the mechanisms involved have yet to be determined.^9^ Based on our data and previous findings, we conclude that patients with DS are not only more likely to be hospitalized with COVID-19, but also that their comorbid conditions, including diabetes in particular, are important drivers.

We also surveyed markers of inflammation, including temperature, C-reactive protein (CRP) concentration, erythrocyte sedimentation rate (ESR), and serum interleukin-1beta (IL-1b), IL-6 and IL-8 concentrations (Supplemental Figure 1A). We found no significant differences between DS patients and controls for any of these markers. Cell counts, including white blood cells, lymphocytes, neutrophils, and eosinophils, were also similar between DS and control patients (Supplemental Figure 1B). Interestingly, the variance of IL-1b and IL-6 concentrations was greater in the DS group, which may indicate that a subset of DS patients had higher levels of soluble inflammatory markers, as previously reported for DS patients in the basal state.^10,11^ Further studies are required to determine whether these cytokines are secreted in larger amounts in DS patients infected with SARS-CoV-2 than in controls, and how they contribute to disease progression in DS.

Finally, patients with DS were more likely to suffer from all the COVID-19-induced complications surveyed, including acute respiratory distress syndrome (ARDS), acute kidney injury (AKI), sepsis, and encephalopathy. Four of the six patients with DS (66.7%) progressed to sepsis, versus only six of the 30 controls (20%) (p<0.05) (Table 2). DS children with sepsis have been reported to have a higher risk of death than non-DS controls with sepsis.^12,13^ This predisposition to COVID-19-induced sepsis is therefore of importance. In addition, hospitalized patients with DS were more likely to receive mechanical ventilation and ICU care, although not significantly so. Hospital stay was longer for patients with DS (median: 17 days, 95% confidence interval [CI]: 14.4-infinity) than for non-DS controls (median: 8 days, CI: 6-15). Two patients in the DS group (33.3%) and two patients in the control group (6.7%) died (Supplemental Figure 1C). The COVID-19 patients with DS hospitalized at Mount Sinai thus had more severe disease progression and longer hospital stays than their non-DS counterparts.

## Discussion

DS is the most common genetic disorder in the US. Currently over 200,000 American have DS. Although medical advances have largely improved the quality of life and longevity of individuals with DS, the immune features of this syndrome remain understudied. Only recently, a few studies have experimentally addressed the perturbed immune state that had been observed in DS. Herein, we report an analysis of individuals with DS who were hospitalized with COVID-19 in the Mount Sinai Health System and discover that patients with DS are at increased risk of COVID-19 hospitalization compared to the general population. While this risk does not appear to be overt, it is prudent to assume a certain level of underestimation of COVID-19 susceptibility in DS due to social behavior. Namely, given lifelong, repeated medical interventions and clinical monitoring, individuals with DS, whether deciding alone or under advice of their caregivers, are likely practicing social isolation more rigorously than the general population. This will only be evident once we are able to evaluate and compare seroprevalence in DS to that of the general population in the same geographic location.

Be that as it may, the six hospitalized patients with DS were on average over ten years younger than non-DS hospitalized patients with COVID-19. However, despite this relatively young age, five out of six of them were above the 86th percentile for age of the DS population. Older age is therefore an important factor for COVID-19 susceptibility in DS as well. Of documented comorbidities associated with COVID-19 susceptibility, diabetes mellitus is likely the most dominant driver of health outcomes in these patients with DS. Of note, we did not find any diagnoses of lower airway anomalies in our patient cohort, although they have been proposed as a cause of increased respiratory infections in patients with DS.^14^ Furthermore, we must also consider the unevaluated parameters which may be contributing to the disease in DS. For example, B cell numbers and quality, T cell function, secreted inflammatory mediators, and response to type I IFNs might all be factors causing an altered response to SARS-CoV-2 infection in DS.

Finally, we found more severe disease progression in DS patients compared to controls. This is consistent with reports that patients with DS are at higher risk of acute lung injury and ARDS compared to controls.^15^ The increased incidence of sepsis in our DS cohort is particularly relevant given studies that patients with DS are at higher risk of mortality secondary to sepsis than septic controls.^12^ Although that study was done in children, the progression to COVID-19-induced sepsis in DS must be closely monitored.

In conclusion, our study highlights that particular attention should be paid to both the prevention and treatment of COVID-19 in individuals with DS, as they are at higher risk of hospitalization induced complications during the SARS-CoV-2 pandemic.

## Data Availability

The authors confirm that the data supporting the findings of this study are available within the article or its supplementary materials.

---

**Supplemental Figure 1.**
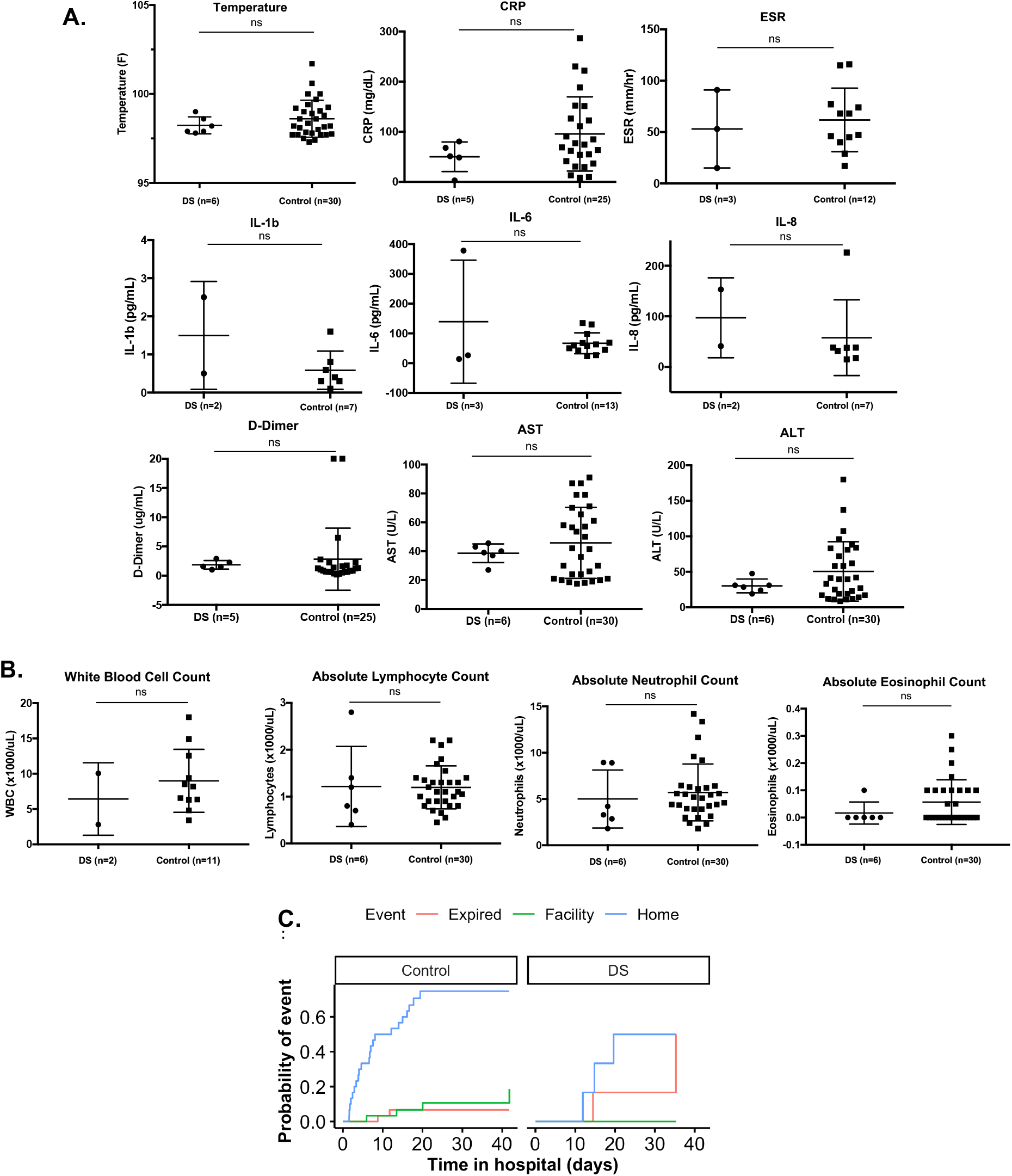
**A**. Median temperature and median laboratory values for C-Reactive Protein (CRP) concentration, Erythrocyte sedimentation rate (ESR), serum Interleukin-1 beta (IL-1 b), IL-6, IL-8, D-dimer, aspartate aminotransferase (AST), and alanine aminotransferase (ALT) concentrations and **B**. Blood leukocytes counts of DS patients and controls hospitalized for COVID-19. **C**. Outcomes of DS and age, sex, and ethnicity-matched controls hospitalized for COVID-19.

